## Supplementary Materials for "Simultaneous estimation of bi-directional causal effects and heritable confounding from GWAS summary statistics"

### Supplementary Materials & Methods

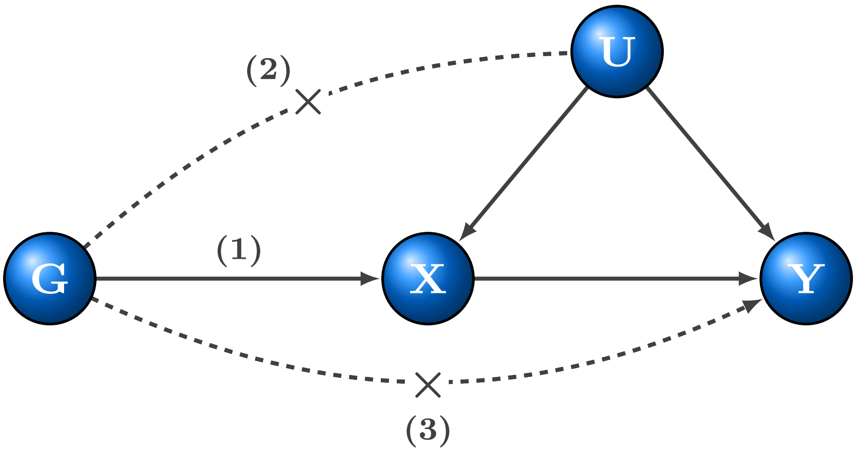


**Figure S1: Basic assumptions of Mendelian randomisation**. (1) Relevance – genetic data, denoted by $G$, is robustly associated with the exposure. (2) Exchangeability – $G$ is not associated with any confounder of the exposure-outcome relationship. (3) Exclusion restriction – $G$ is independent of the outcome conditional on the exposure and all confounders of the exposure-outcome relationship (i.e. the only path between the instrument and the outcome is via the exposure).

#### 1.1 Likelihood function identifiability

The likelihood function is symmetric around $U$, but for simplicity we will consider the general case where the variables of $U$ and $X$ are flipped, although the same can be said for the variables of $U$ and $Y$. The likelihood function is partially identifiable such that there exists for any given model parameters, another model with different parameters but with the exact same likelihood function.

Proof: given that the SNPs effects between trait $X$ and the confounder $U$ are flipped, the new parameters follow the following structure:

$$
\begin{aligned}
h'_x &= t_x + t_y \cdot \alpha_{y\to x} \\
h'_y &= h_y \\
\alpha'_{y\to x} &= \alpha_{y\to x} \\
\alpha'_{x\to y} &= \frac{q_x \cdot \alpha_{x\to y} + q_y}{q_x + q_y \cdot \alpha_{y\to x}} \\
&= \frac{q_x(\alpha_{x\to y} + \frac{q_y}{q_x})}{q_x(1 + \frac{q_y}{q_x} \cdot \alpha_{y\to x})} \\
&= \frac{\alpha_{x\to y} + \frac{q_y}{q_x}}{1 + \frac{q_y}{q_x} \cdot \alpha_{y\to x}}
\end{aligned}
$$

through inverse transformation,

$$
\alpha_{x\to y} = \frac{\alpha'_{x\to y} + \frac{q'_y}{q'_x}}{1 + \frac{q'_y}{q'_x} \cdot \alpha_{y\to x}}
$$

Plugging in $\alpha'_{x\to y}$ in the above equation, and simplifying $\frac{ty}{tx}$ by $w$ and $\frac{ty'}{tx'}$ by $w'$ to get the confounding ratio:

$$
\begin{aligned}
\alpha_{x\to y} &= \frac{\alpha'_{x\to y} + w'}{1 + w' \cdot \alpha_{y\to x}} \\
\alpha_{x\to y} + \alpha_{x\to y} \cdot w' \cdot \alpha_{y\to x} &= \alpha'_{x\to y} + w' \\
\alpha_{x\to y} - \alpha'_{x\to y} &= w' - \alpha_{x\to y} \cdot w' \cdot \alpha_{y\to x} \\
\alpha_{x\to y} - \alpha'_{x\to y} &= w'(1 - \alpha_{x\to y} \cdot \alpha_{y\to x}) \\
w' &= \frac{\alpha_{x\to y} - \alpha'_{x\to y}}{1 - \alpha_{x\to y} \cdot \alpha_{y\to x}}
\end{aligned}
$$

inserting the complete form of $\alpha'_{x\to y}$,

$$\begin{aligned}
w' &= \frac{\alpha_{x\to y} - \frac{\alpha_{x\to y} + \frac{q_y}{q_x}}{1+\frac{q_y}{q_x}\cdot\alpha_{y\to x}}}{1-\alpha_{x\to y}\cdot\alpha_{y\to x}} \\
&= \frac{\alpha_{x\to y}(1+w\cdot\alpha_{y\to x}) - w - \alpha_{x\to y}}{(1-\alpha_{x\to y}\cdot\alpha_{y\to x})(1+w\cdot\alpha_{y\to x})} \\
&= \frac{\alpha_{x\to y}\cdot w\cdot\alpha_{y\to x} - w}{(1-\alpha_{x\to y}\cdot\alpha_{y\to x})(1+w\cdot\alpha_{y\to x})} \\
&= \frac{w(\alpha_{x\to y}\cdot\alpha_{y\to x} - 1)}{(1-\alpha_{x\to y}\cdot\alpha_{y\to x})(1+w\cdot\alpha_{y\to x})} \\
&= \frac{-w}{1+w\cdot\alpha_{y\to x}}
\end{aligned}$$

In order to obtain $t'_y$ and $t'_x$, we use the equations of $h'_x$, $\alpha'_{x\to y}$ and by using the inverse transformation of $\alpha'_{y\to x} = \alpha_{y\to x}$, $\alpha_{x\to y}$ as well as $w'$ as follows:

$$\begin{aligned}
t'_y &= \frac{-t'_x\cdot w}{1+w\cdot\alpha_{y\to x}} \\
h_x &= t'_x + t'_y\cdot\alpha_{y\to x} \\
&= t'_x + \frac{-t'_x\cdot w}{1+w\cdot\alpha_{y\to x}}\cdot\alpha_{y\to x} \\
&= \frac{t'_x + t'_x\cdot w\cdot\alpha_{y\to x} - t'_x\cdot w\cdot\alpha_{y\to x}}{1+w\cdot\alpha_{y\to x}} \\
&= \frac{t'_x}{1+w\cdot\alpha_{y\to x}} \\
t'_x &= h_x(1+w\cdot\alpha_{y\to x})
\end{aligned}$$

Replacing $t'_x$ in $h_x$ to get $t'_y$:

$$t'_y = h_x\cdot w$$

Under these two models with equal likelihood, there are three slopes obtained from the observed data: two are the correlation of effect sizes ($\alpha_{x\to y}$ and $1/\alpha_{y\to x}$), where one of them is greater than, and the other is within the parameter bounds. The third is the correlation of the confounder $\frac{\alpha_{x\to y}+\frac{q_y}{q_x}}{1+\frac{q_y}{q_x}\cdot\alpha_{y\to x}}$.

More often than not, only one slope is recovered within the boundaries of the parameters set for LHC-MR. However, given the now known re-parameterisation, the second (and if found, third) slope can be simply calculated if not found by the likelihood function minimisation. It is reasonable to assume that the direct heritability of each trait is larger than the indirect heritability, hence we report parameter sets where $h_x^2 > t_x^2$ or $h_y^2 > t_y^2$.

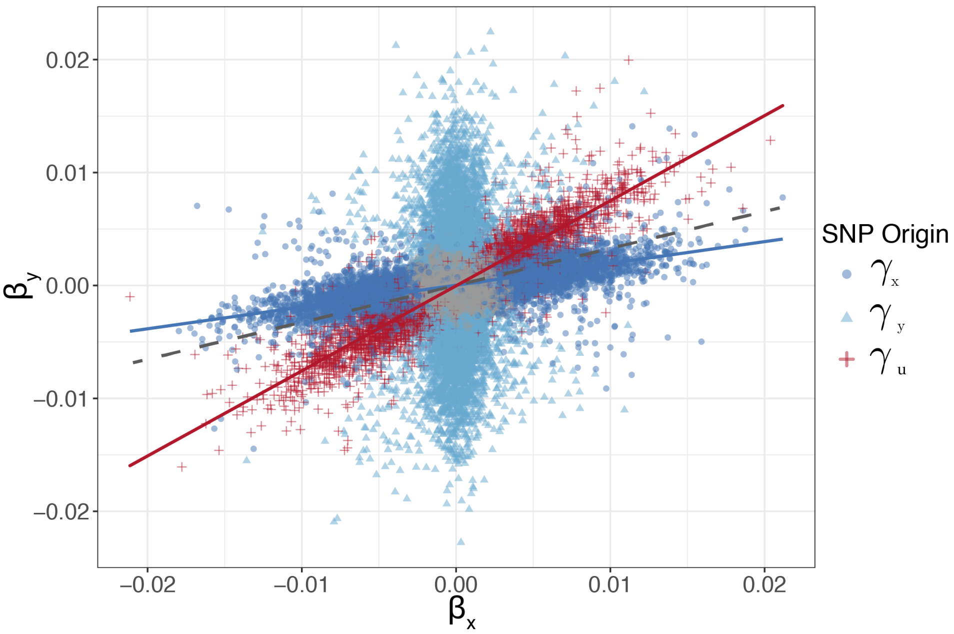


**Figure S2: An illustration of a scatter plot showing simulated observed SNP effects on traits $X$ and $Y$, coloured by the strongest effect between the three vectors** $\gamma_x, \gamma_y, \gamma_u$. SNPs in grey are those with no effect on any of the traits. This illustration shows the distinct clusters that could arise in the presence of a confounder. The dark blue cluster of SNPs represents those that are not in violation of any of the MR assumption, and hence its slope reflects the true causal effect of $X$ on $Y$, while the red cluster of SNPs are those associated with the confounder. The steeper slope of the red cluster of SNPs causes a typical regression line - shown in grey - that represents the causal effect (estimated using conventional MR methods) to be overestimated.

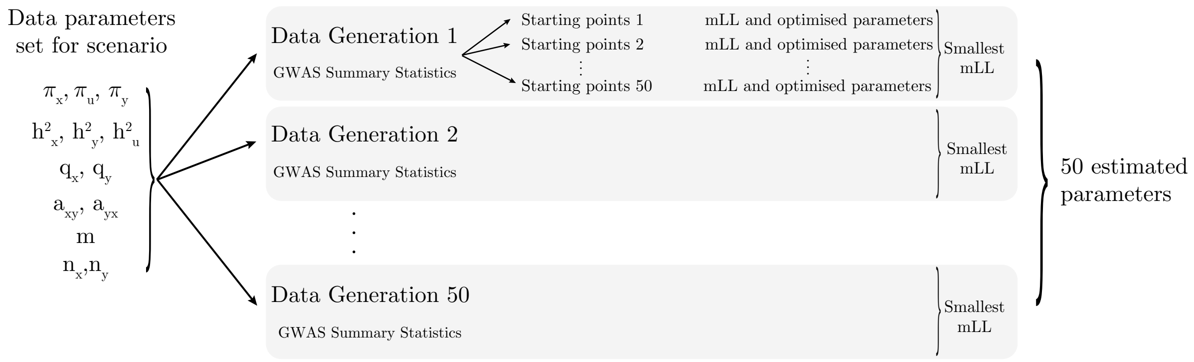


**Figure S3: A schema showing the workflow of the simulation results**. For a single set of parameter settings, 50 different data generations of GWAS summary statistics are created for trait $X$ and $Y$. The summary statistics of a single data generation, as well as the sample size, SNP number and SNP-based LD structure are used in the likelihood optimisation function that is run with 100 different random starting points in order to explore the likelihood surface. A single maximum likelihood and its corresponding estimated parameters are selected to represent the estimates of that data generation. And this is repeated for the other generations. The results for several data generation are often represented in boxplots throughout the paper.

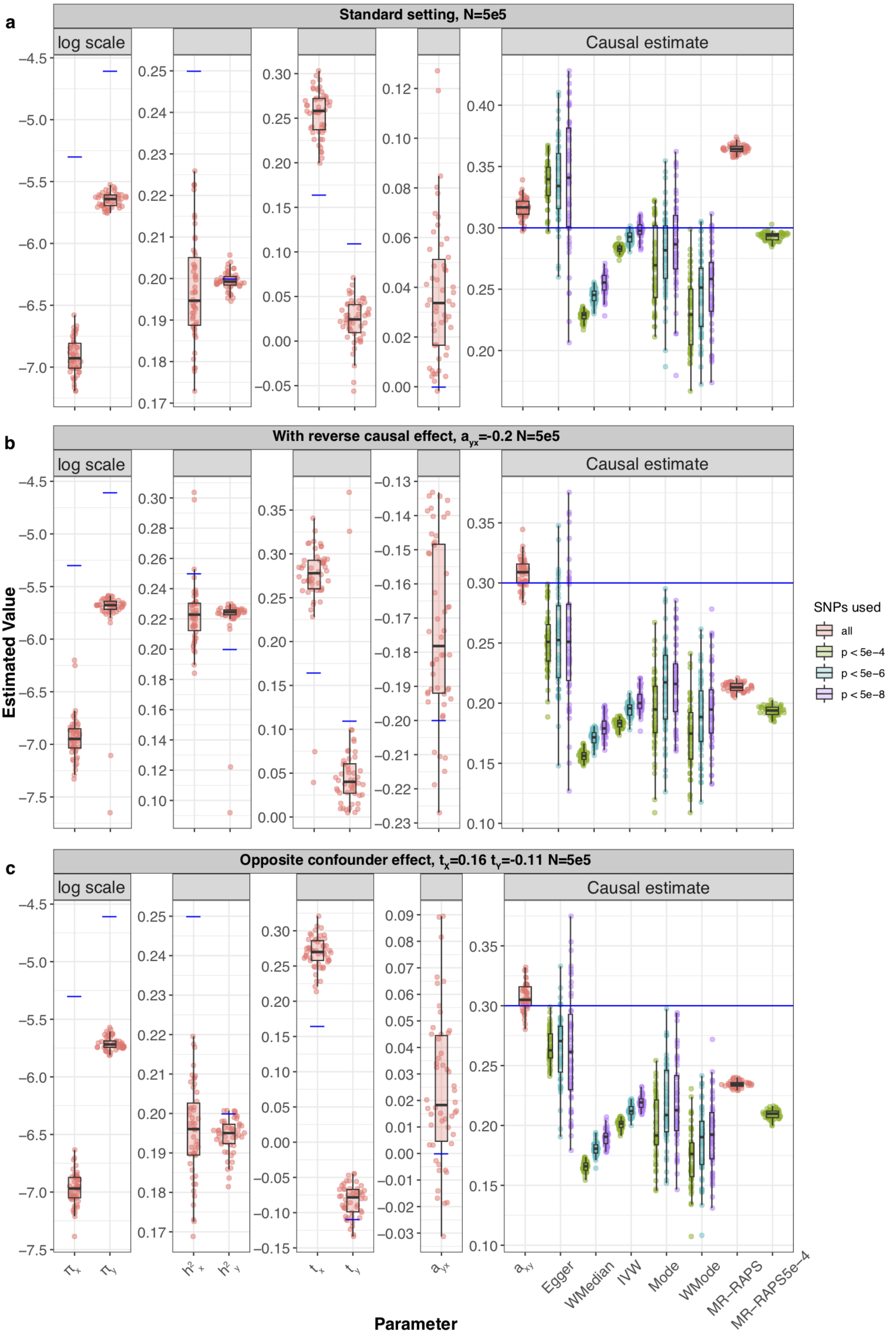


**Figure S4: Simulation results under various scenarios.** These Raincloud boxplots[28] represent the distribution of parameter estimates from 50 different data generations under various conditions. For each generation, standard MR methods as well as our LHC-MR were used to estimate a causal effect. The true values of the parameters used in the data generations are represented by the blue dots/lines. **a** Estimation under standard settings ($\pi_x = 5 \times 10^{-3}, \pi_y = 1 \times 10^{-2}, \pi_u = 5 \times 10^{-2}, h_x^2 = 0.25, h_y^2 = 0.2, h_u^2 = 0.3, t_x = 0.16, t_y = 0.11$). **b** Addition of a reverse causal effect $\alpha_{y\to x} = -0.2$. **c** Confounder with opposite causal effects on $X$ and $Y$ ($t_x = 0.16, t_y = -0.11$).

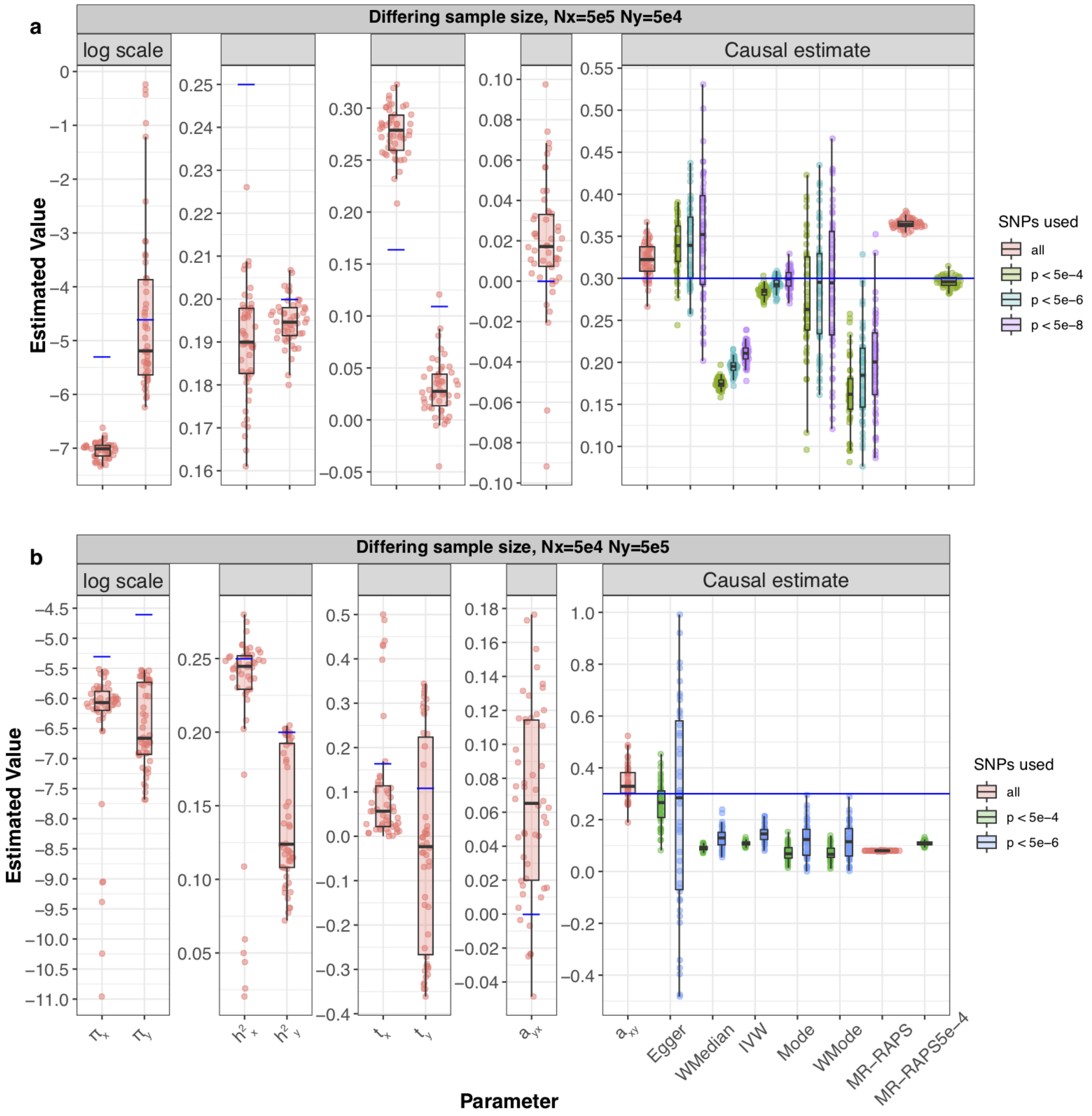


**Figure S5: Simulation results showing varying sample sizes for the two exposure and outcome samples**. Raincloud boxplots representing the distribution of parameter estimates from 50 different data generations. For each generation, standard MR methods as well as our LHC-MR were used to estimate a causal effect. The true values of the parameters used in the data generations are represented by the blue dots/lines. In this figure, samples sizes for the two traits differ as such $n_x = 500{,}000$ and $n_y = 50{,}000$ for **a**, and $n_x = 50{,}000$ and $n_y = 500{,}000$ for **b**.

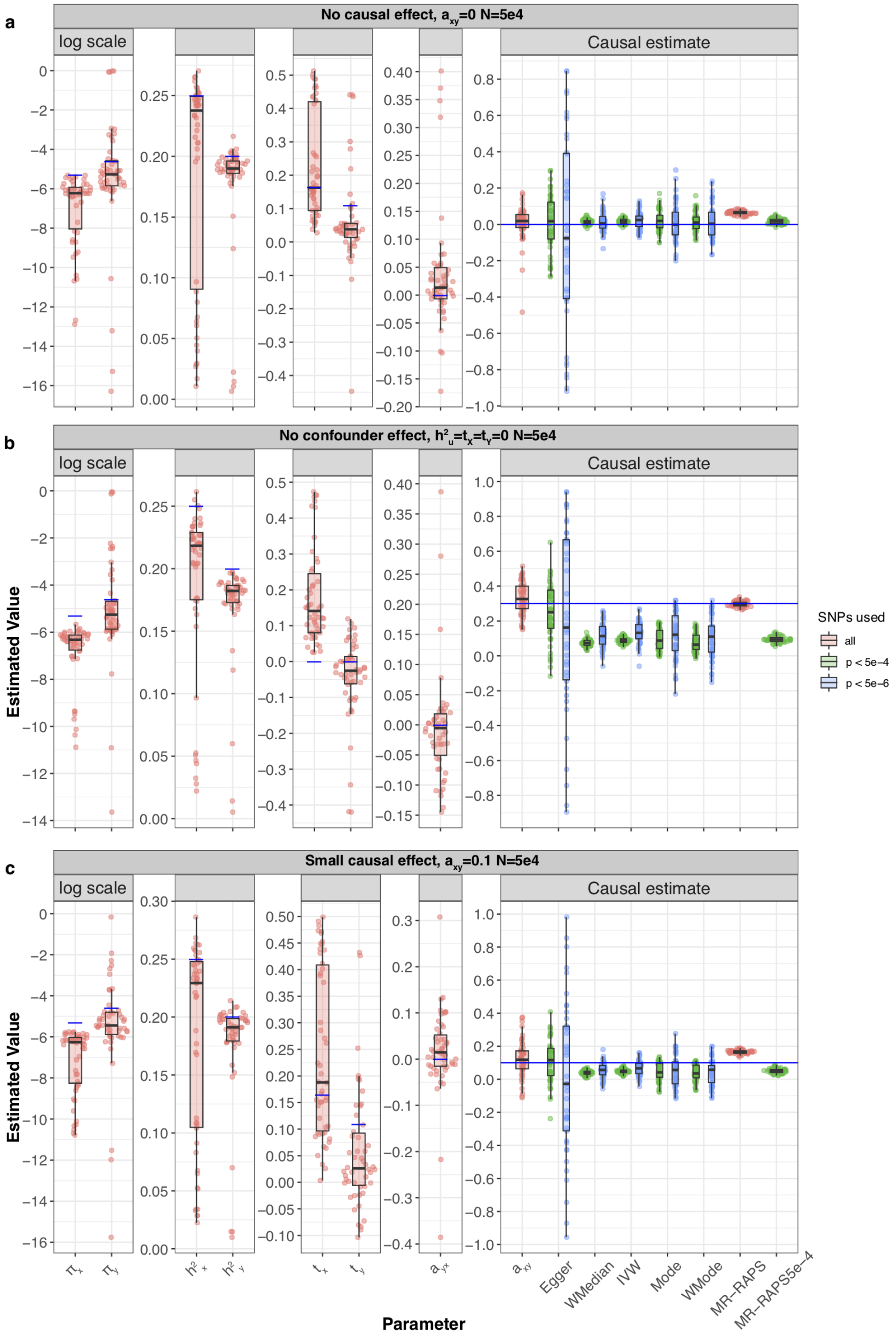


**Figure S6: Simulation results under various scenarios.** These Raincloud boxplots[28] represent the distribution of parameter estimates from 50 different data generations under various conditions. For each generation, standard MR methods as well as our LHC-MR were used to estimate a causal effect. The true values of the parameters used in the data generations are represented by the blue dots/lines. **a** The data simulated had no causal effect in either direction. **b** The data simulated had no confounder effect with $\pi_u, t_x$, and $t_y = 0$. **c** This model had a small causal effect of $\alpha_{x\rightarrow y} = 0.1$.

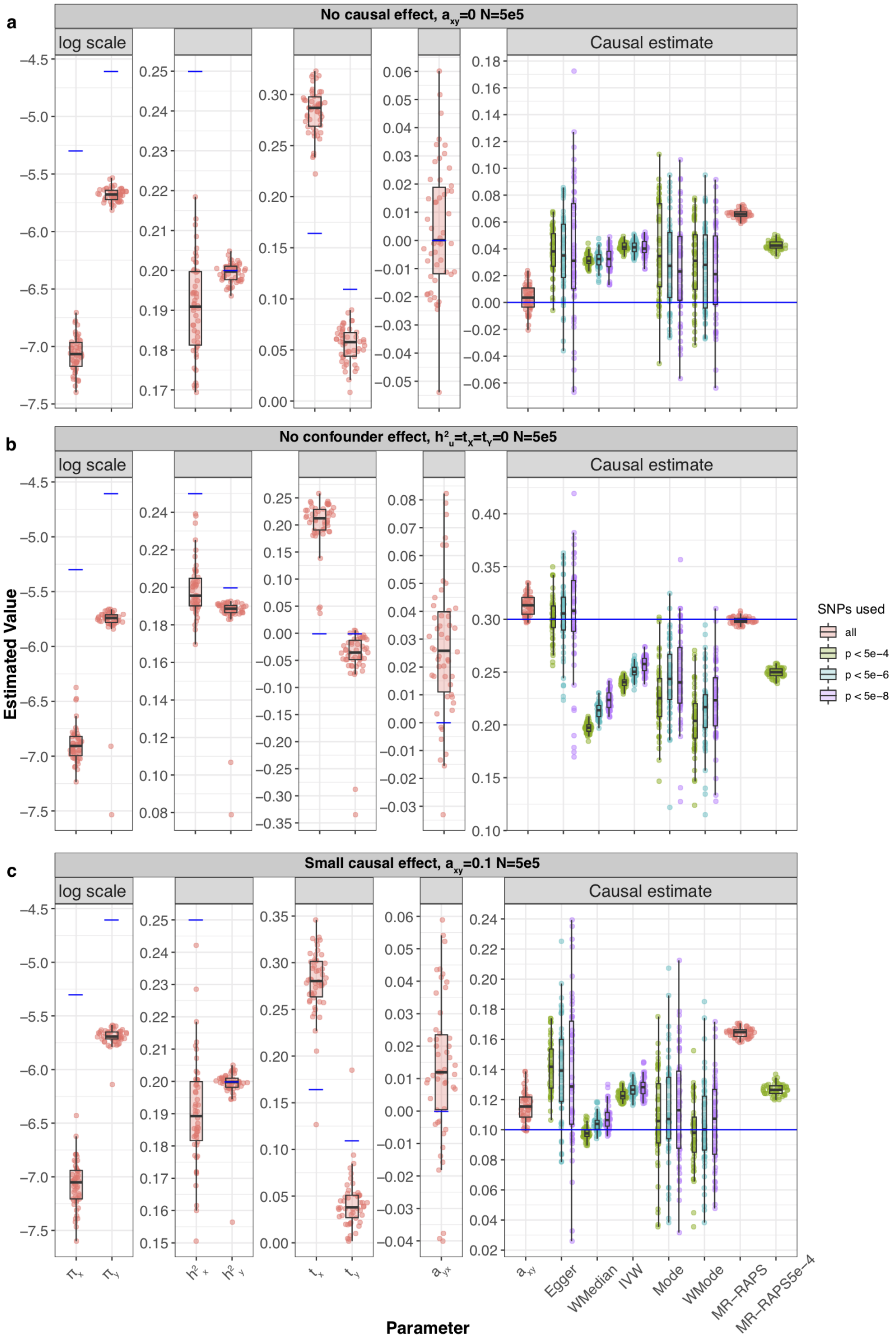


**Figure S7: Simulation results under various scenarios.** These Raincloud boxplots[28] represent the distribution of parameter estimates from 50 different data generations under various conditions. For each generation, standard MR methods as well as our LHC-MR were used to estimate a causal effect. The true values of the parameters used in the data generations are represented by the blue dots/lines. **a** The data simulated had no causal effect in either direction. **b** The data simulated had no confounder effect with $\pi_u, t_x$, and $t_y = 0$. **c** This model had a small causal effect of $\alpha_{x \to y} = 0.1$.

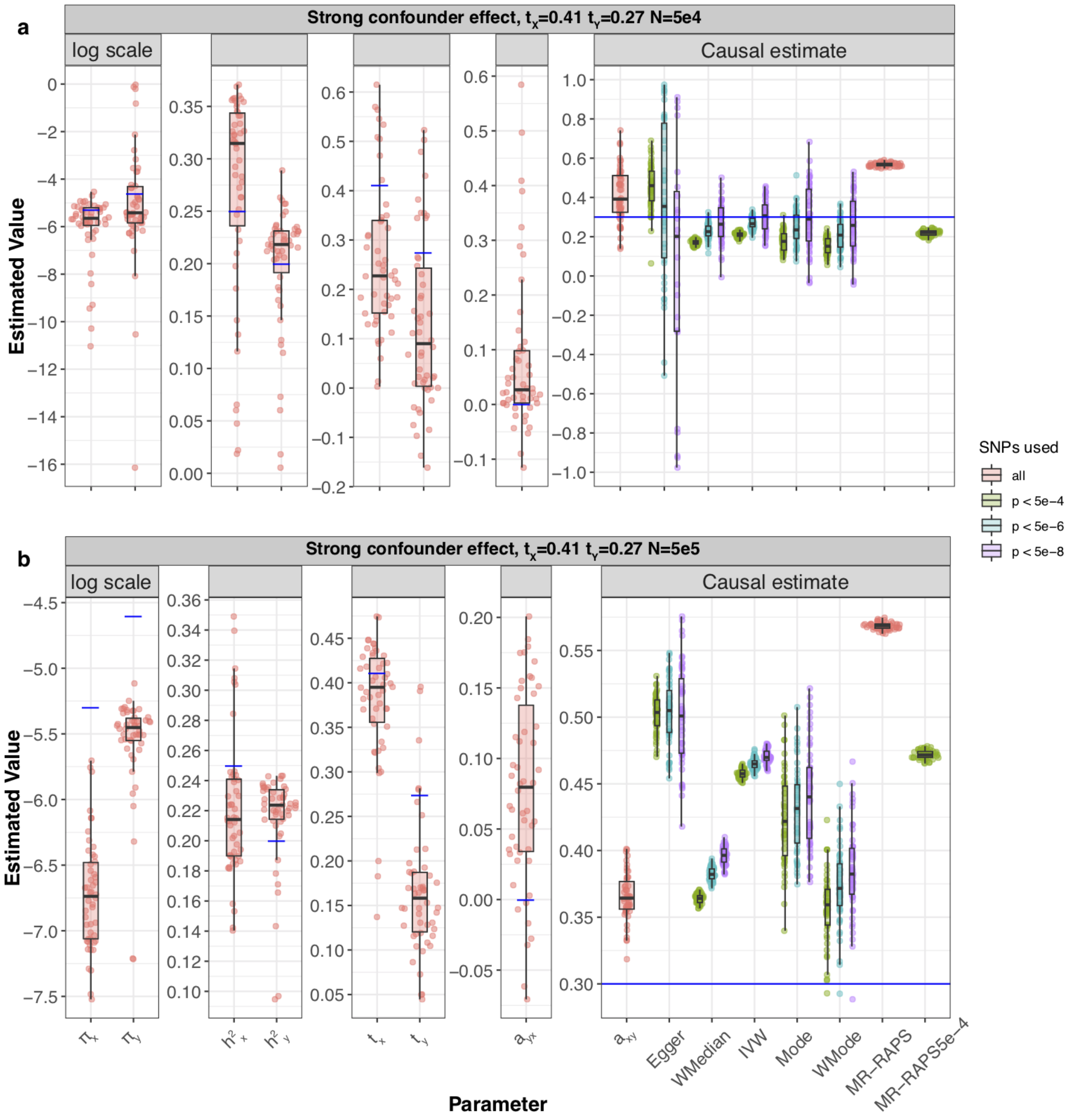


**Figure S8: Simulation results under various scenarios.** These Raincloud boxplots[28] represent the distribution of parameter estimates from 50 different data generations under various conditions. For each generation, standard MR methods as well as our LHC-MR were used to estimate a causal effect. The true values of the parameters used in the data generations are represented by the blue dots/lines. **a** The data simulated shows the increased effect of $U$ on $X$ and $Y$ through $t_x = 0.41, t_y = 0.27$ instead of the standard setting $t_x = 0.16, t_y = 0.11$. **b** This panel show the same thing but with a larger sample size of $n_x = n_y = 500,000$

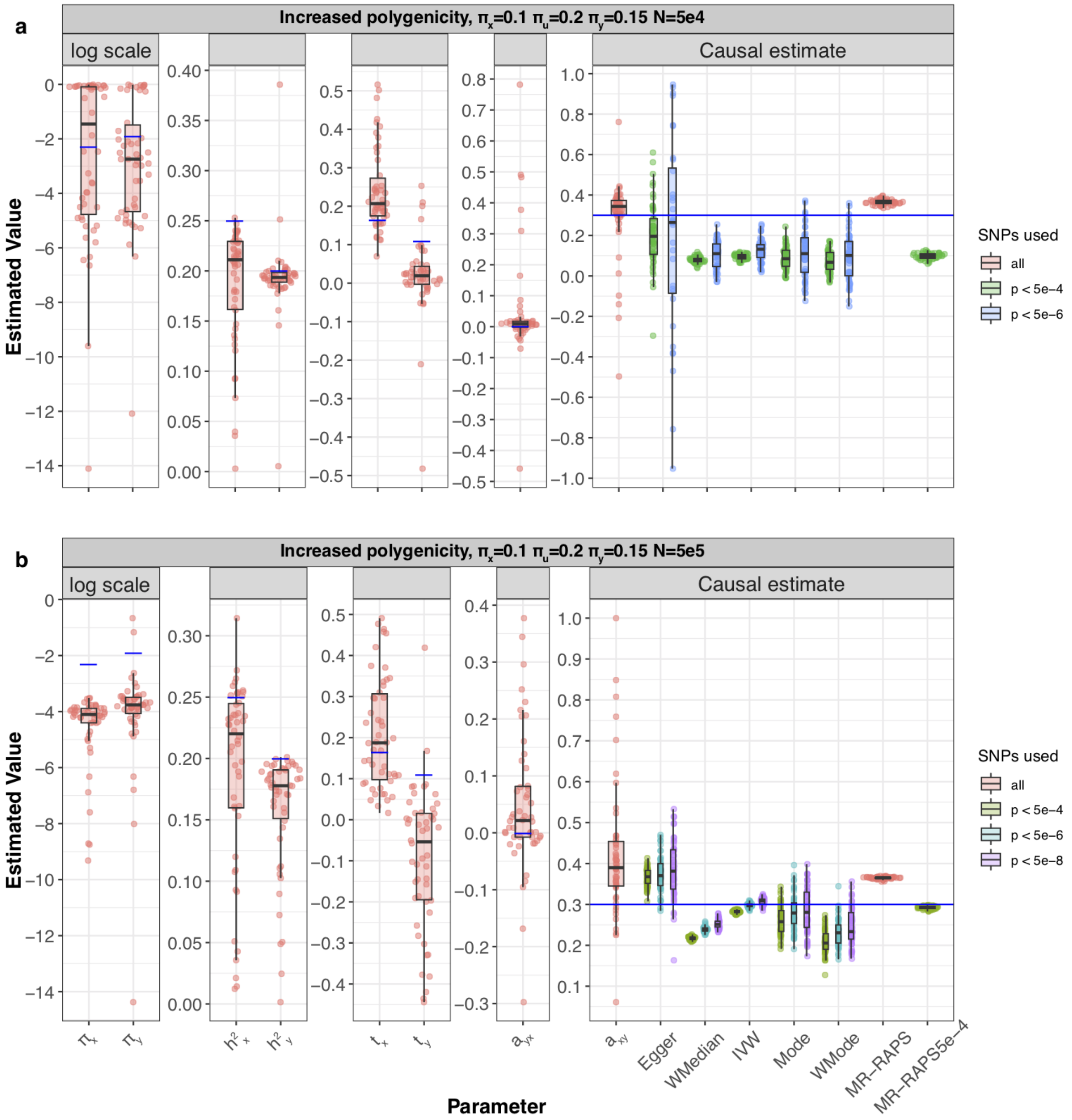


**Figure S9: Simulation results where there is an increased polygenicity for all traits.** Boxplots representing the distribution of parameter estimates from 100 different data generations. For each generation, standard MR methods as well as our LHC-MR were used to estimate a causal effect. The true values of the parameters used in the data generations are represented by the blue dots/lines. The proportion of effective SNPs that make up the spike-and-slab distributions of the $\gamma$ vectors in this setting is $10\%, 15\%, and 20\%$ for traits $X, Y$ and $U$ respectively. **a** Results for smaller sample size of $n_x = n_y = 50,000$. **b** Results for larger sample size of $n_x = n_y = 500,000$.

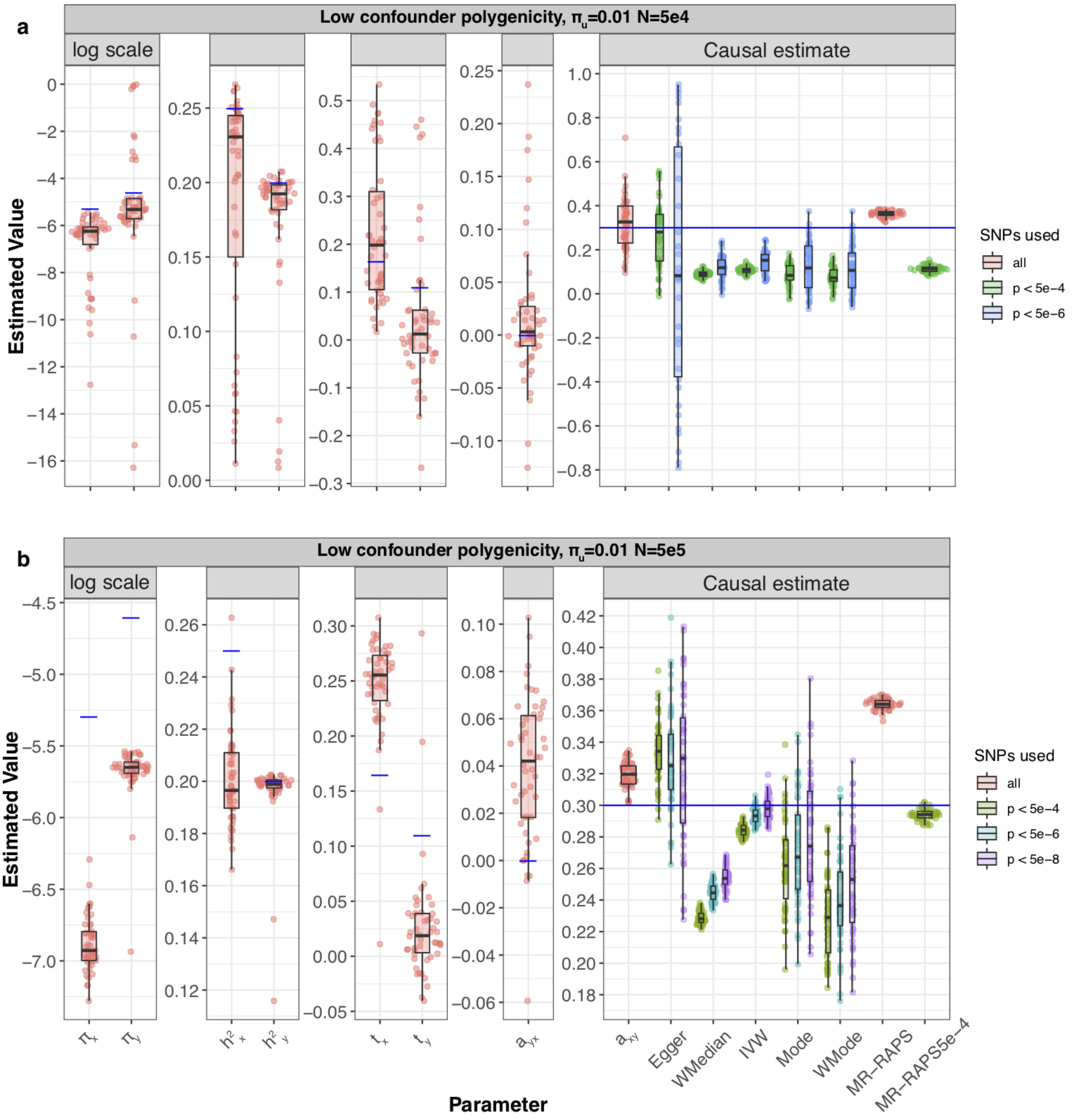


**Figure S10: Simulation results where the polygenicity of the confounder is reduced.** Boxplots representing the distribution of parameter estimates from 100 different data generations. For each generation, standard MR methods as well as our LHC-MR were used to estimate a causal effect. The true values of the parameters used in the data generations are represented by the blue dots/lines. In this figure, the polygenicity for $U$ is decreased in the form of lower $\pi_u = 0.01$. **a** Results for smaller sample size of $n_x = n_y = 50,000$. **b** Results for larger sample size of $n_x = n_y = 500,000$.

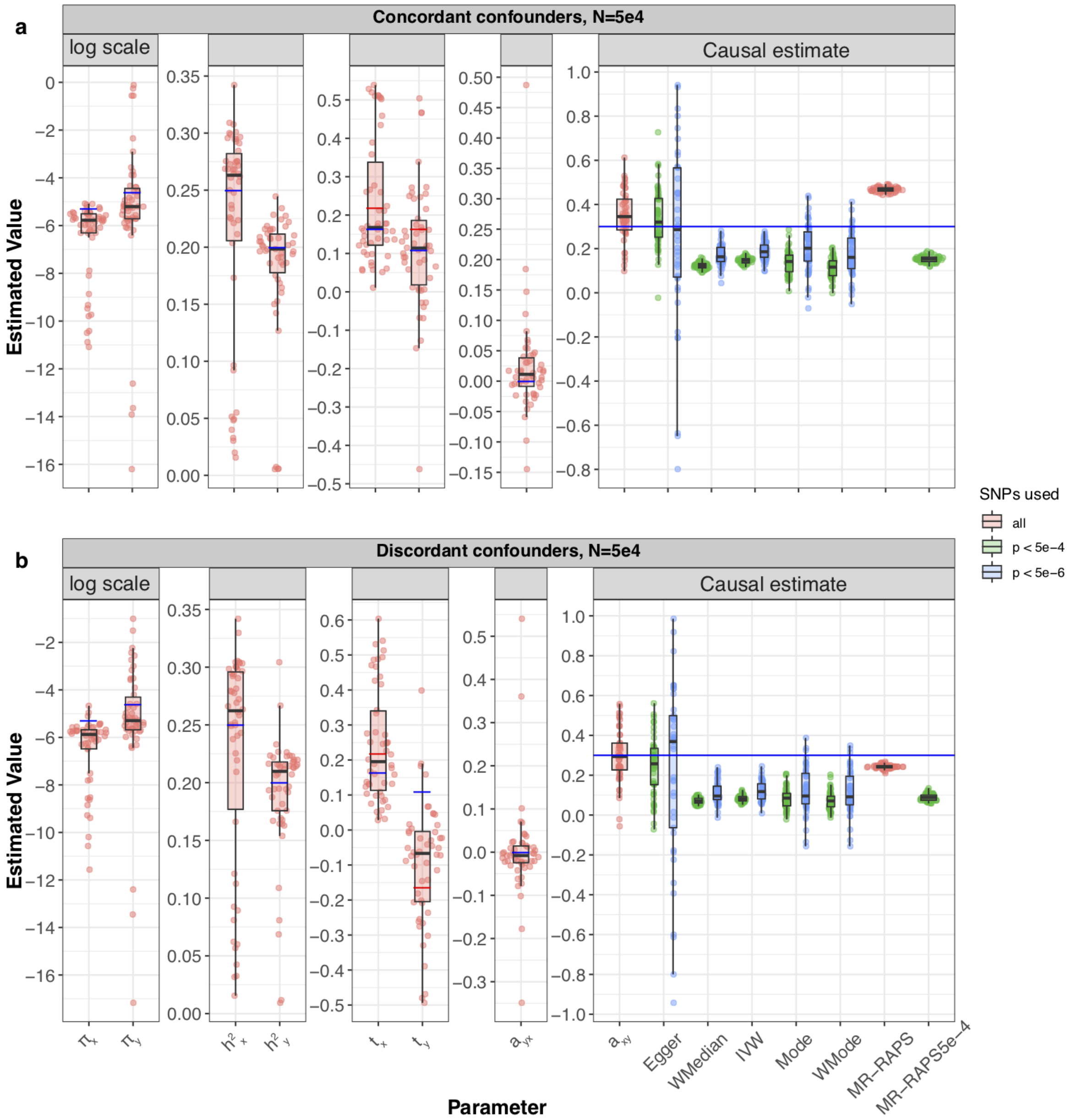


**Figure S11: Simulation results where there are two underlying confounders, once with concordant and another with discordant effects on the exposure-outcome pair.** Boxplots representing the distribution of parameter estimates from 100 different data generations. For each generation, standard MR methods as well as our LHC-MR were used to estimate a causal effect. The true values of the parameters used in the data generations are represented by the blue dots/lines. **a** The underlying data generations have two concordant heritable confounders $U_1$ and $U_2$ with positive effects on traits $X$ and $Y$. **b** The data generations have two discordant heritable confounders with $t_x^{(1)} = 0.16, t_y^{(1)} = 0.11$ shown as blue dots and $t_x^{(2)} = 0.22, t_y^{(2)} = -0.16$ shown as red dots.

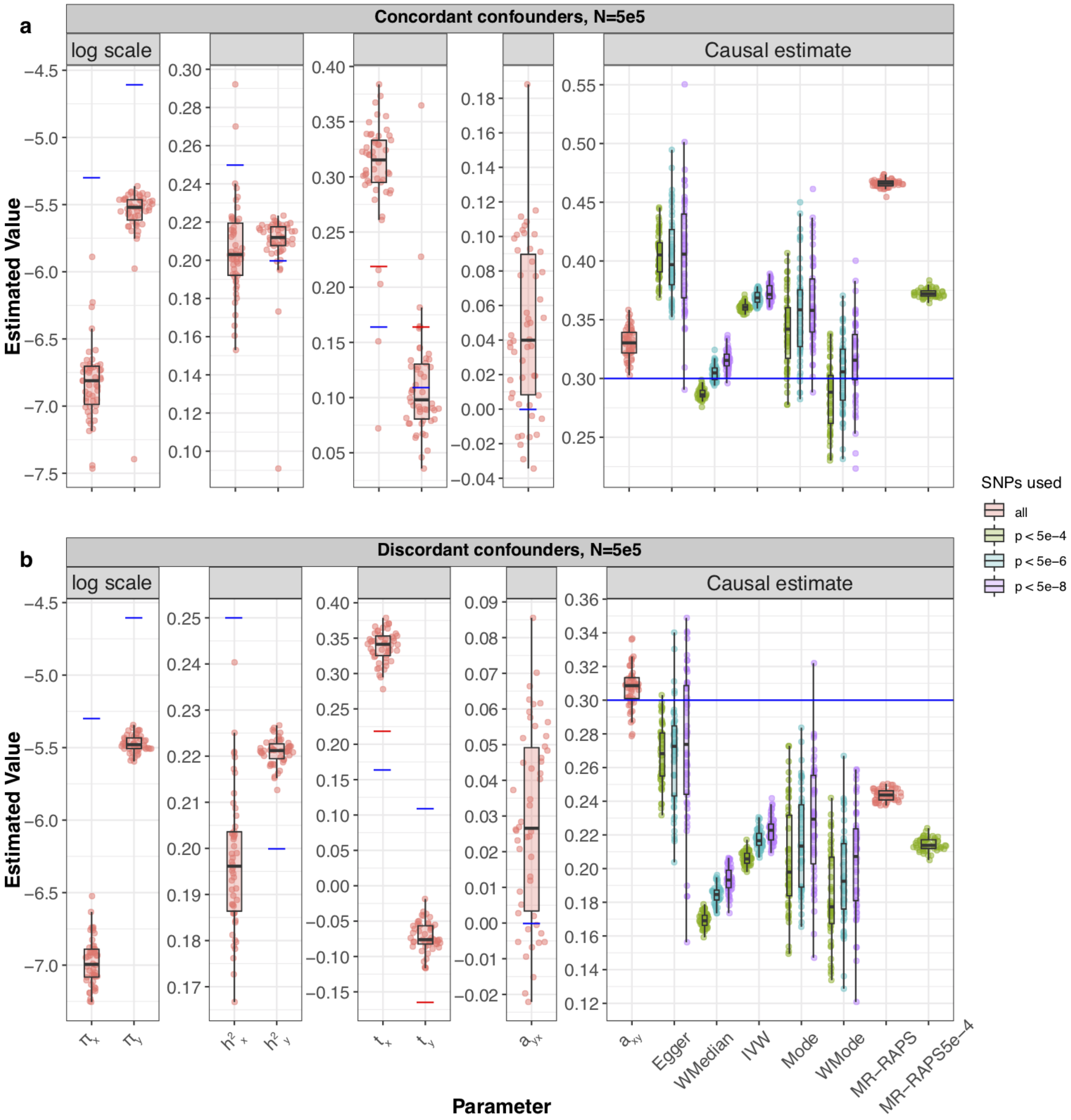


**Figure S12: Simulation results where there are two underlying confounders, once with concordant and another with discordant effects on the exposure-outcome pair.** Boxplots representing the distribution of parameter estimates from 100 different data generations. For each generation, standard MR methods as well as our LHC-MR were used to estimate a causal effect. The true values of the parameters used in the data generations are represented by the blue dots/lines. **a** The underlying data generations have two concordant heritable confounders $U_1$ and $U_2$ with positive effects on traits $X$ and $Y$. **b** The data generations have two discordant heritable confounders with $t_x^{(1)} = 0.16, t_y^{(1)} = 0.11$ shown as blue dots and $t_x^{(2)} = 0.22, t_y^{(2)} = -0.16$ shown as red dots.

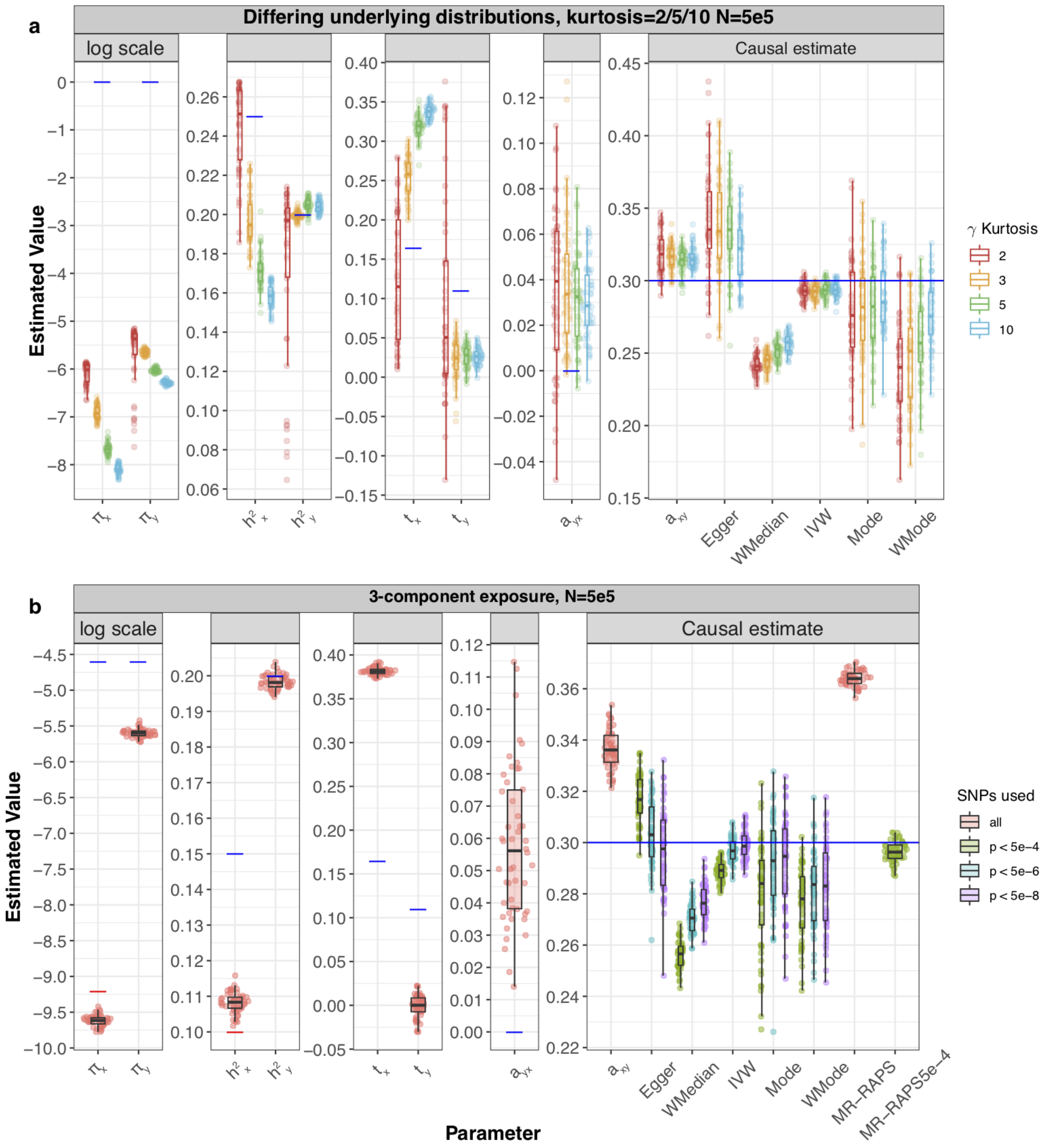


**Figure S13: Simulation results under various scenarios.** These Raincloud boxplots[28] represent the distribution of parameter estimates from 50 different data generations under various conditions. For each generation, standard MR methods as well as our LHC-MR were used to estimate a causal effect. The true values of the parameters used in the data generations are represented by the blue dots/lines. **a** The different coloured boxplots represent the underlying non-normal distribution used in the simulation of the three $\gamma_x, \gamma_x, \gamma_u$ vectors associated to their respective traits. The Pearson distributions had the same 0 mean and skewness, however their kurtosis ranged between 2 and 10, including the kurtosis of 3, which corresponds to a normal distribution assumed by our model. The standard MR results reported had IVs selected with a p-value threshold of $5 \times 10^{-6}$. **b** Addition of a third component for exposure $X$, while decreasing the strength of $U$. True parameter values are in colour, blue and red for each component ($\pi_{x1} = 1 \times 10^{-4}, \pi_{x2} = 1 \times 10^{-2}, h^2_{x1} = 0.15, h^2_{x2} = 0.1$).

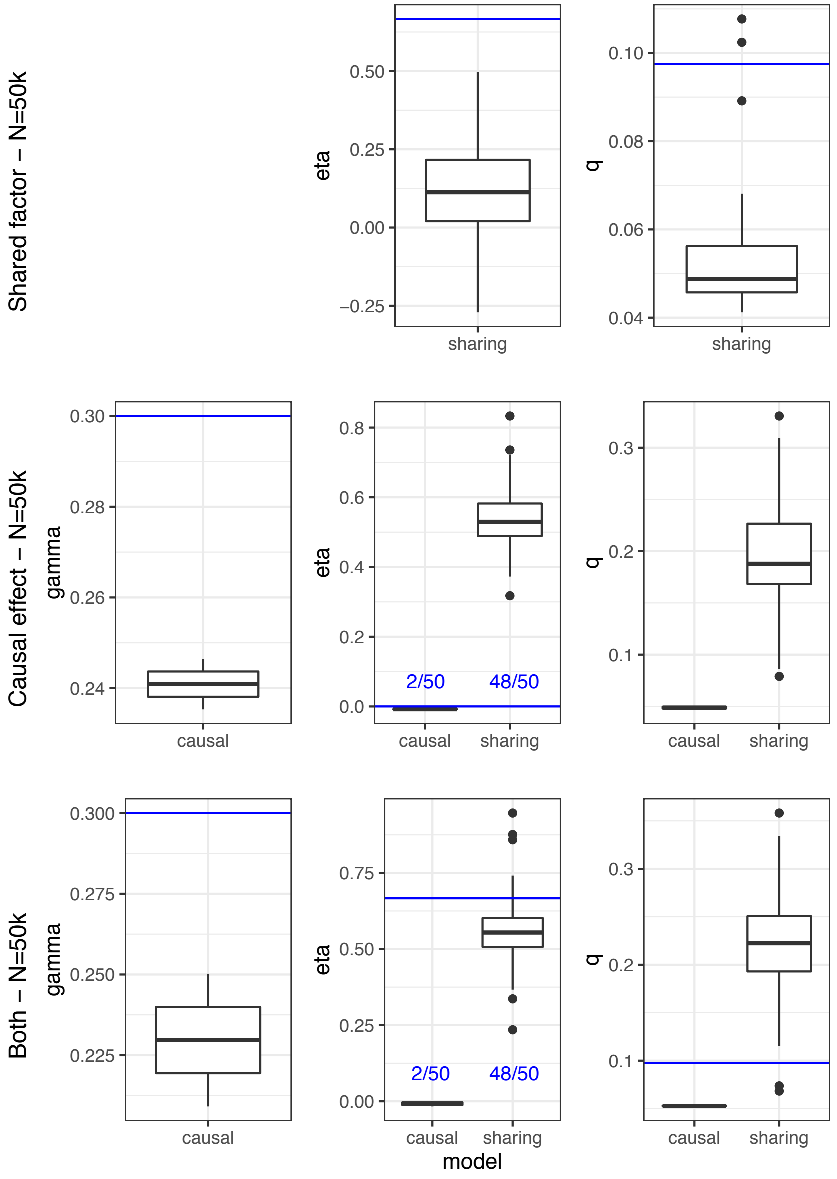


**Figure S14: Running CAUSE on LHC-MR simulated data under the standard settings**. Boxplots of the parameter estimation of CAUSE on LHC-simulated data ($n_x = n_y = 50,000$) under three different scenarios: presence of a shared factor only, presence of a causal effect only, presence of both. CAUSE returns two possible models with a respective p-value, the sharing and the causal model, where the causal mode is the significant of the two. When only an underlying shared factor was present in the simulated data, CAUSE had no significant causal estimates. With a true underlying causal effect, or when both an underlying causal effect and a shared factor was present, the causal model was significant only 4% of the simulations.

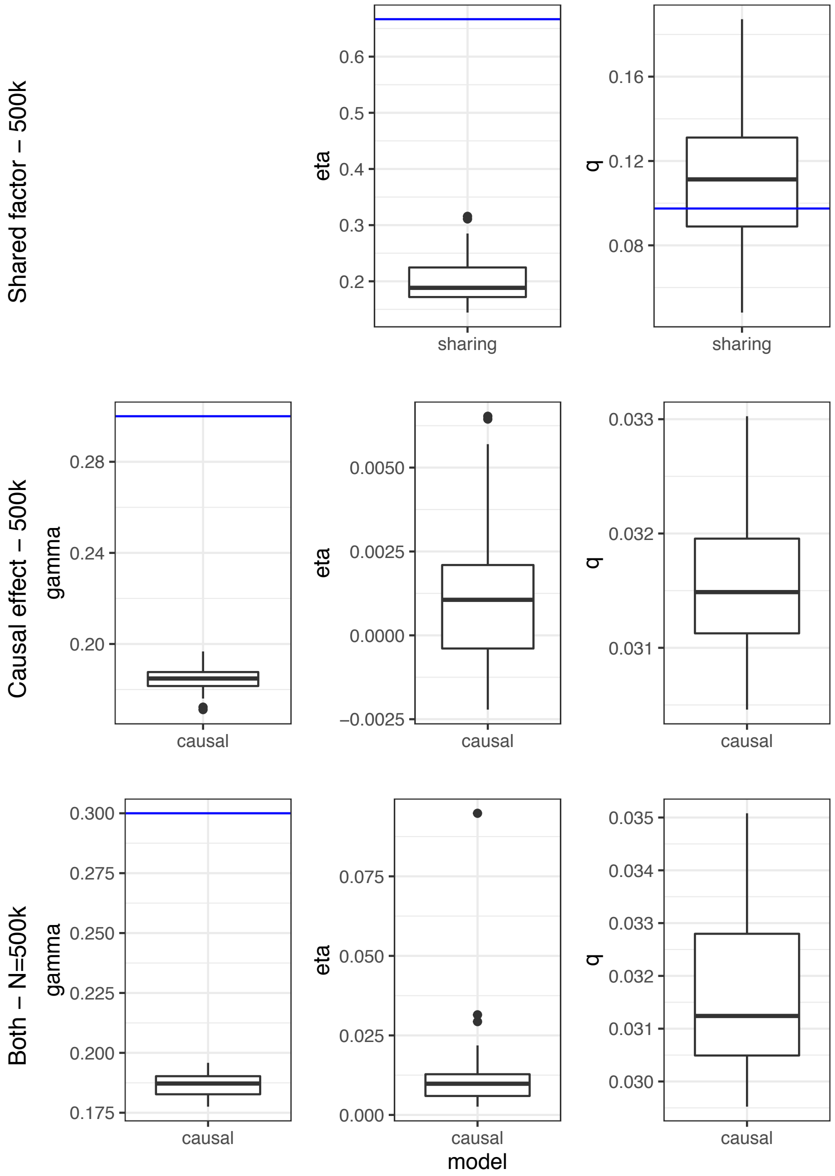


**Figure S15: Running CAUSE on LHC-MR simulated data under the standard settings**. Boxplots of the parameter estimation of CAUSE on LHC-simulated data ($n_x = n_y = 500,000$) under three different scenarios: presence of a shared factor only, presence of a causal effect only, presence of both. CAUSE returns two possible models with a respective p-value, the sharing and the causal model, where the causal mode is the significant of the two. When only an underlying shared factor was present in the simulated data, CAUSE had no significant causal estimates. With a true underlying causal effect, or when both an underlying causal effect and a shared factor was present, the causal model was significant 100% of the simulations.

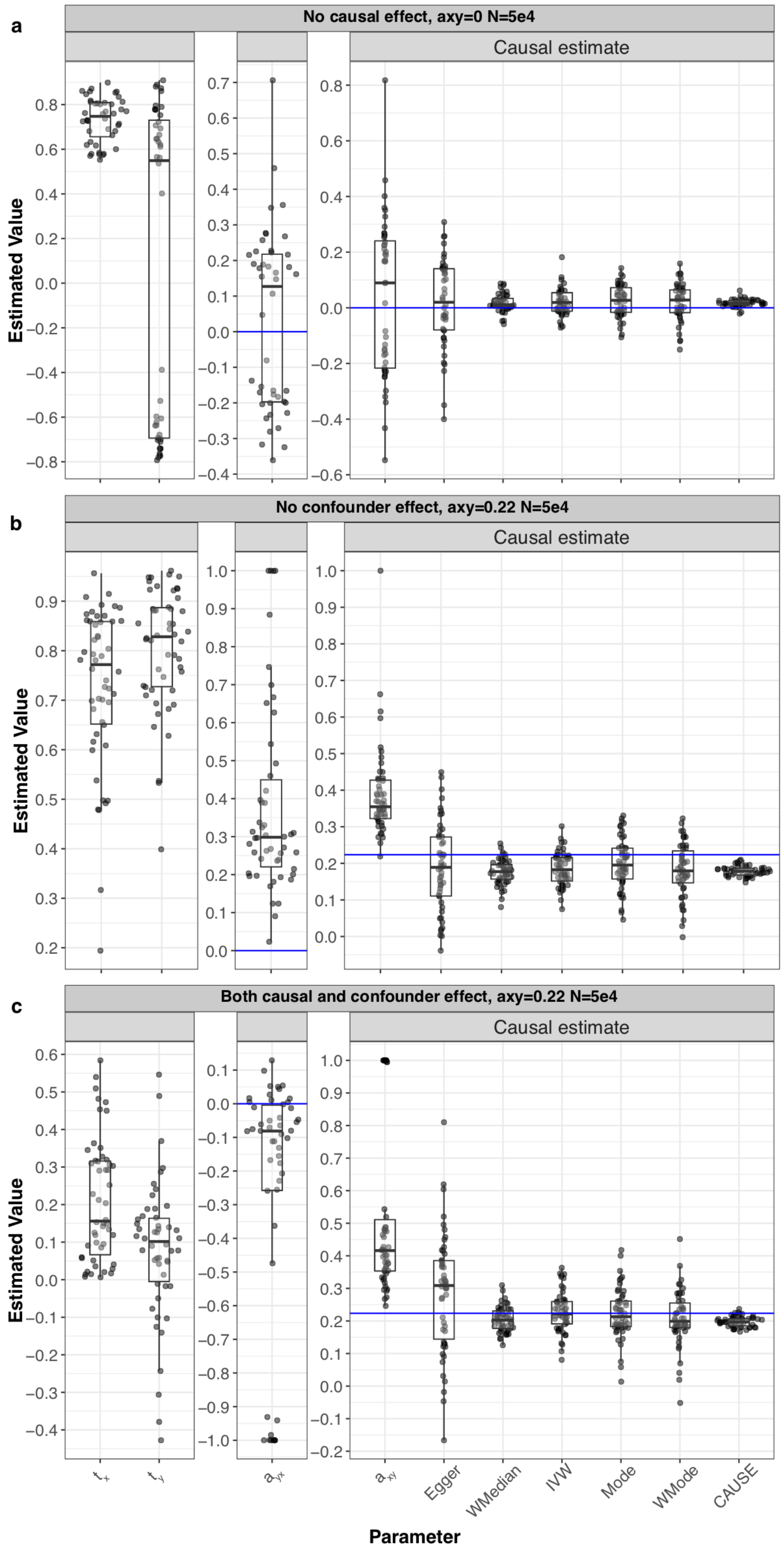


**Figure S16: Running LHC-MR on CAUSE simulated data under various scenarios**. Raincloud boxplots representing the distribution of parameter estimates from LHC-MR of 50 different data generations using the CAUSE framework. For each generation, standard MR methods, CAUSE as well as our LHC-MR were used to estimate a causal effect. The true values of the parameters used in the data generations are represented by the blue dots/lines. **a** CAUSE data was generated with no causal effect but with a shared factor with an $\eta$ value of $\sim 0.22$. CAUSE chooses a sharing model 100% of the time with no estimate for a causal effect. **b** CAUSE is simulated with causal effect but with no shared factor. **c** CAUSE is simulated with both a causal effect and a shared factor.

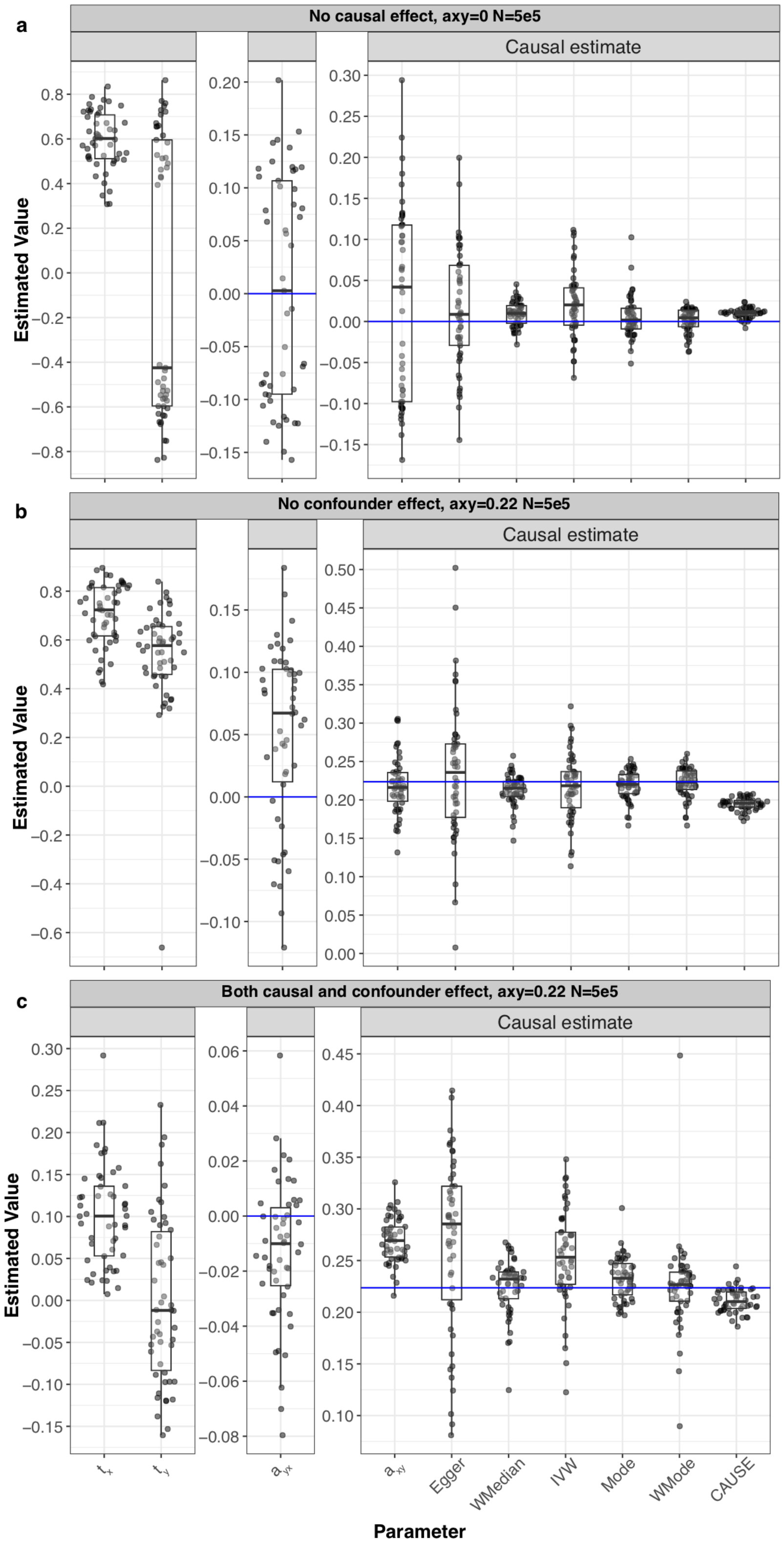


**Figure S17: Running LHC-MR on CAUSE simulated data under various scenarios**. Raincloud boxplots representing the distribution of parameter estimates from LHC-MR of 50 different data generations using the CAUSE framework. For each generation, standard MR methods, CAUSE as well as our LHC-MR were used to estimate a causal effect. The true values of the parameters used in the data generations are represented by the blue dots/lines. **a** CAUSE data was generated with no causal effect but with a shared factor with an $\eta$ value of $\sim 0.22$. **b** CAUSE is simulated with causal effect but with no shared factor. **c** CAUSE is simulated with both a causal effect and a shared factor. LHC-MR seems to exhibit a bimodal effect at first glance, but the two peaks are not connected.

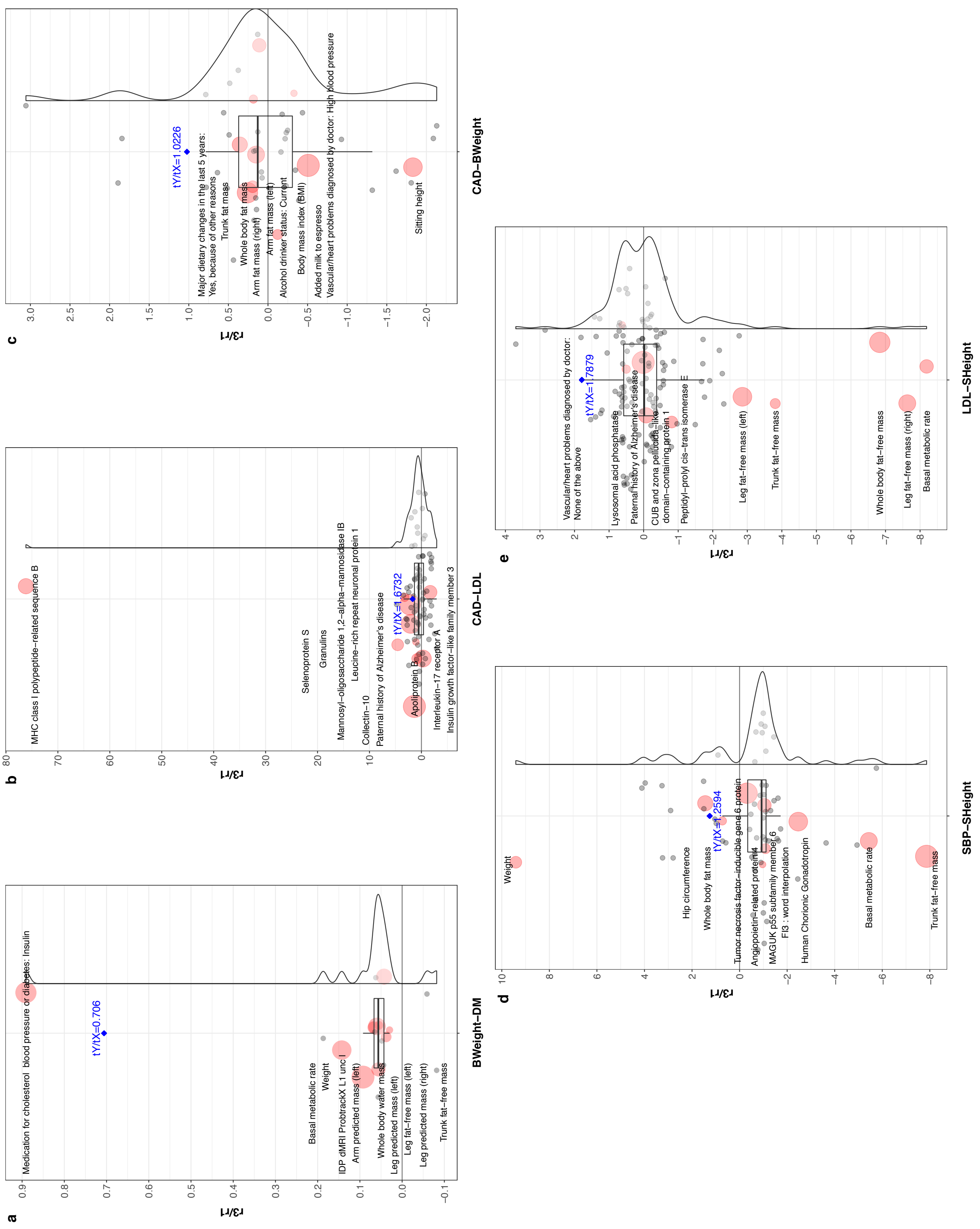

a
BWeight–DM
r3/r1
tY/tX=0.706
Medication for cholesterol blood pressure or diabetes: Insulin
Basal metabolic rate
Weight
IDP dMRI ProbtrackX L1 unc l
Arm predicted mass (left)
Whole body water mass
Leg predicted mass (left)
Leg fat–free mass (left)
Leg predicted mass (right)
Trunk fat–free mass
b
CAD–LDL
r3/r1
tY/tX=1.6732
MHC class I polypeptide–related sequence B
Selenoprotein S
Granulins
Mannosyl–oligosaccharide 1,2–alpha–mannosidase IB
Leucine–rich repeat neuronal protein 1
Collectin–10
Paternal history of Alzheimer's disease
Apolipoprotein B
Interleukin–17 receptor A
Insulin growth factor–like family member 3
c
CAD–BWeight
r3/r1
tY/tX=1.0226
Major dietary changes in the last 5 years:
Yes, because of other reasons
Trunk fat mass
Whole body fat mass
Arm fat mass (right)
Arm fat mass (left)
Alcohol drinker status: Current
Body mass index (BMI)
Added milk to espresso
Vascular/heart problems diagnosed by doctor: High blood pressure
Sitting height
d
SBP–SHeight
r3/r1
tY/tX=1.2594
Weight
Hip circumference
Whole body fat mass
Tumor necrosis factor–inducible gene 6 protein
Angiopoietin–related protein 4
MAGUK p55 subfamily member 6
FI3 : word interpolation
Human Chorionic Gonadotropin
Basal metabolic rate
Trunk fat–free mass
e
LDL–SHeight
r3/r1
tY/tX=1.7879
Vascular/heart problems diagnosed by doctor:
None of the above
Lysosomal acid phosphatase
Paternal history of Alzheimer's disease
CUB and zona pellucida–like
domain–containing protein 1
Peptidyl–prolyl cis–trans isomerase E
Leg fat–free mass (left)
Trunk fat–free mass
Whole body fat–free mass
Leg fat–free mass (right)
Basal metabolic rate

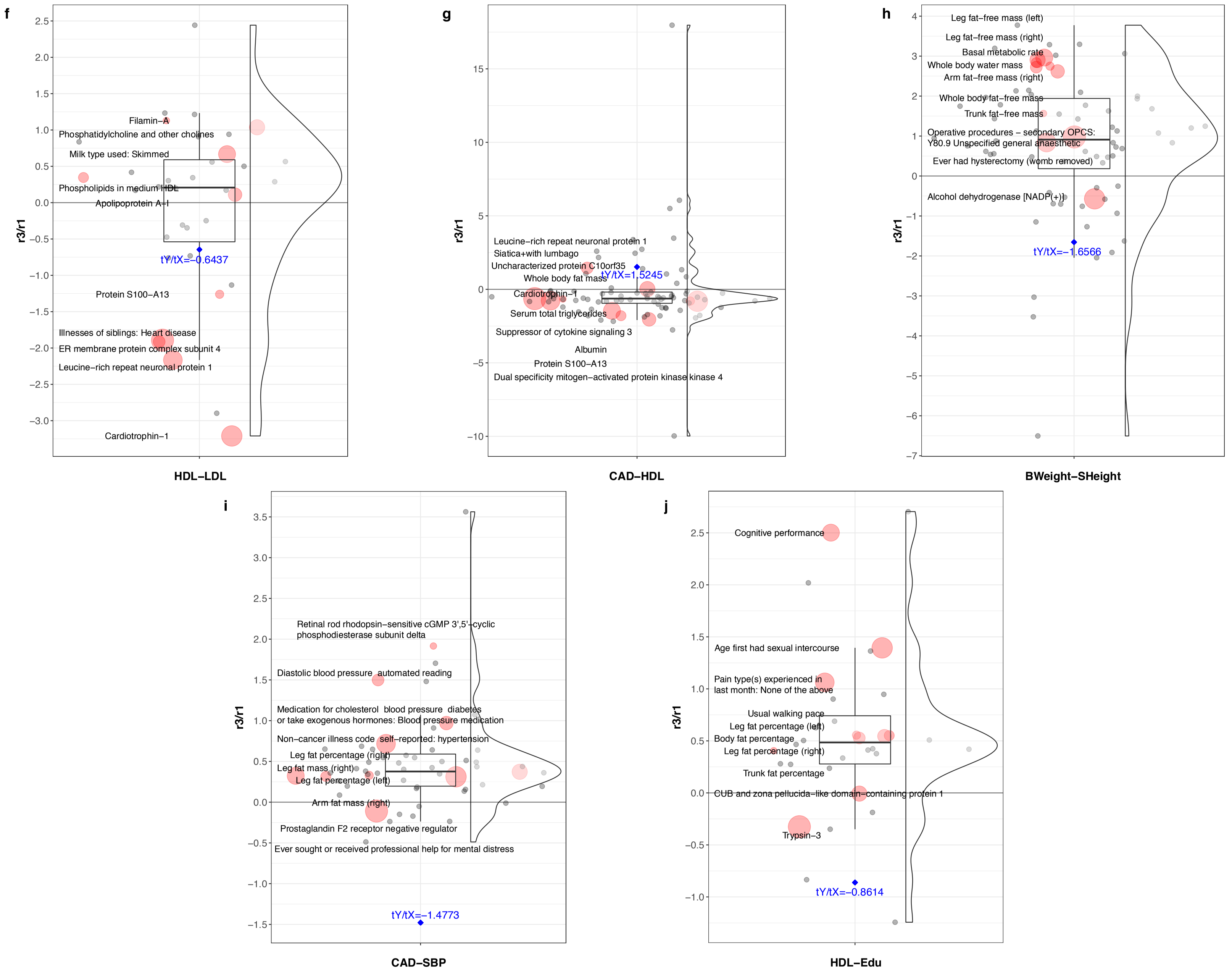


**Figure S18: Confounder effects obtained from EpiGraphDB plotted as a raincloud with the $r_3/r_1$ ratio on the y-axis**. The blue diamonds represent the $t_y/t_x$ ratio derived from the LHC model for that trait pair, also reported in blue text. Labelled dots in red and their varying size show the ten largest confounder traits in terms of their absolute effect product on the two traits, whereas grey dots represent the rest of the confounder traits found by EpigraphDB.

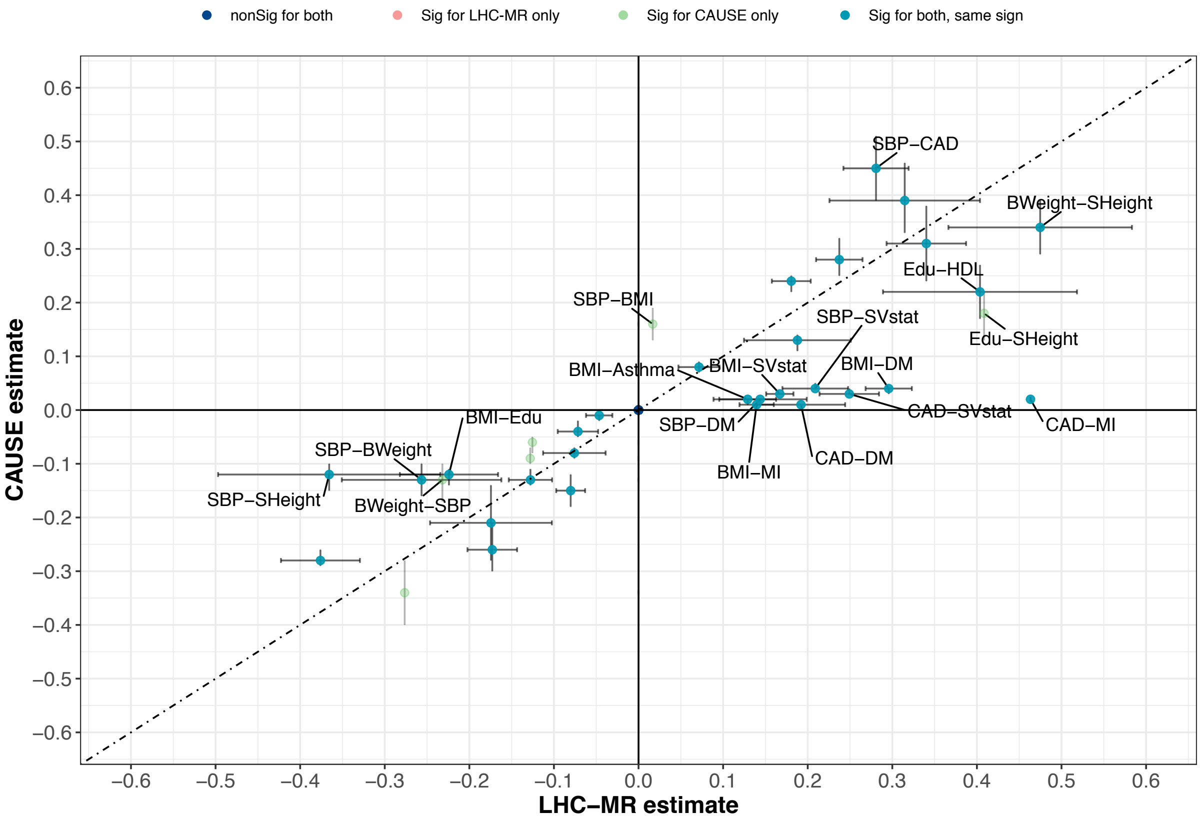


**Figure S19: A scatter plot of the causal effect estimates between LHC-MR and CAUSE.** To improve visibility, non-significant estimates by both methods are placed at the origin, while significant estimates by both methods appear on the diagonal with 95% CI error bars for LHC-MR estimates, and 95% credible interval error bars for CAUSE estimates. Labelled pairs are those with an estimate difference greater than 0.1.

### Supplementary Tables

| UKBB ID / Data Origin | Trait Name | Abbreviation | Sample Size | PMID |
|---|---|---|---|---|
| **845** | Age completed full time education | Edu | 240,547 | 25826379 |
| **21001_irnt** | Body mass index (BMI) | BMI | 359,983 | 25826379 |
| **2443** | Diabetes diagnosed by doctor | DM | 360,192 | 25826379 |
| **20002_1075** | Non-cancer illness code, self-reported: heart attack/myocardial infarction | MI | 361,141 | 25826379 |
| **20002_1111** | Non-cancer illness code, self-reported: asthma | Asthma | 361,141 | 25826379 |
| **2887** | Number of cigarettes previously smoked daily | PSmoke | 84,456 | 25826379 |
| **20022_irnt** | Birth weight | BWeight | 205,475 | 25826379 |
| **50_irnt** | Standing height | SHeight | 360,388 | 25826379 |
| **4080** | Systolic blood pressure, automated reading | SBP | 340,159 | 25826379 |
| **20003_1140861958** | Treatment/medication code: simvastatin | SVstat | 361,141 | 25826379 |
| **30780_irnt** | LDL Cholesterol | LDL | 343,621 | 25826379 |
| **30760_irnt** | HDL Cholesterol | HDL | 315,133 | 25826379 |
| **UKBB + CARDIoGRAMplusC4D** | Coronary Artery Disease | CAD | 380,831 | 29212778 |

**Table S1: Details of the origin study of each trait, its abbreviation used in this paper, the sample size of the study for that trait, as well as the PubMed article ID.**

**a** — **MR-Egger** (rows: LHC-MR)

| LHC-MR | Sig+ | Sig− | nonSig |
|---|---|---|---|
| Sig+ | 10 | 0 | 27 |
| Sig− | 0 | 2 | 35 |
| nonSig | 0 | 2 | 56 |

**b** — **WMedian**

| | Sig+ | Sig− | nonSig |
|---|---|---|---|
| Sig+ | 21 | 1 | 15 |
| Sig− | 0 | 12 | 25 |
| nonSig | 1 | 6 | 51 |

**c** — **IVW**

| | Sig+ | Sig− | nonSig |
|---|---|---|---|
| Sig+ | 23 | 0 | 14 |
| Sig− | 0 | 17 | 20 |
| nonSig | 2 | 5 | 51 |

**d** — **Mode** (rows: LHC-MR)

| LHC-MR | Sig+ | Sig− | nonSig |
|---|---|---|---|
| Sig+ | 12 | 0 | 25 |
| Sig− | 0 | 2 | 35 |
| nonSig | 0 | 0 | 58 |

**e** — **WMode**

| | Sig+ | Sig− | nonSig |
|---|---|---|---|
| Sig+ | 13 | 3 | 21 |
| Sig− | 0 | 7 | 30 |
| nonSig | 0 | 2 | 56 |

**f** — **WMode** (rows: MR-Egger)

| MR-Egger | Sig+ | Sig− | nonSig |
|---|---|---|---|
| Sig+ | 10 | 0 | 0 |
| Sig− | 0 | 1 | 3 |
| nonSig | 3 | 11 | 104 |

**Table S7: Cross tables between LHC-MR and various standard MR methods comparing the significance and sign of each respective causal estimate.** **f** shows a cross table between the two-least correlated MR methods in terms of their estimates.

| Pair | $\alpha_{x\to y}$ | p-value | $\gamma$ | IVW $\alpha_{x\to y}$ | p-value |
|---|---|---|---|---|---|
| BMI-Asthma | 0.1290 | 4.99E-14 | 0.02 (0.01, 0.02) | 0.0593 | 1.00E-08 |
| BMI-DM | 0.2958 | 1.07E-99 | 0.04 (0.03, 0.04) | 0.2447 | 2.25E-140 |
| BMI-SBP | 0.1878 | 5.55E-09 | 0.13 (0.11, 0.14) | 0.1547 | 1.11E-24 |
| BMI-SVstat | 0.1670 | 2.08E-91 | 0.03 (0.03, 0.03) | 0.1570 | 4.26E-63 |
| BMI-MI | 0.1396 | 1.67E-41 | 0.01 (0.01, 0.01) | 0.1027 | 9.16E-32 |
| BWeight-SHeight | 0.4748 | 9.60E-18 | 0.34 (0.29, 0.39) | 0.2959 | 8.01E-10 |
| SHeight-BWeight | 0.1806 | 1.93E-53 | 0.24 (0.22, 0.25) | 0.1803 | 7.21E-86 |
| SBP-DM | 0.1437 | 3.17E-07 | 0.02 (0.01, 0.02) | 0.0697 | 3.69E-07 |
| DM-SVstat | 0.3147 | 4.11E-12 | 0.39 (0.33, 0.46) | 0.2524 | 1.28E-16 |
| SHeight-Edu | 0.0715 | 8.42E-09 | 0.08 (0.07, 0.09) | 0.0643 | 2.28E-21 |
| SBP-SVstat | 0.2089 | 4.84E-26 | 0.04 (0.04, 0.05) | 0.1853 | 1.46E-52 |
| Edu-HDL | 0.4037 | 5.25E-12 | 0.22 (0.17, 0.27) | 0.2848 | 4.06E-08 |
| BMI-CAD | 0.2373 | 2.37E-64 | 0.28 (0.25, 0.32) | 0.1800 | 2.42E-53 |
| CAD-DM | 0.1920 | 5.92E-13 | 0.01 (0.01, 0.01) | 0.0659 | 0.002455431 |
| DM-CAD | 0.4283 | 5.60E-19 | 1.95 (1.26, 2.64) | 0.1796 | 4.15E-05 |
| SBP-CAD | 0.2807 | 2.86E-46 | 0.45 (0.39, 0.51) | 0.2500 | 9.77E-24 |
| CAD-SVstat | 0.2491 | 8.82E-44 | 0.03 (0.03, 0.04) | 0.3077 | 1.15E-25 |
| CAD-MI | 0.4634 | 0 | 0.02 (0.02, 0.02) | 0.4191 | 3.07E-285 |
| LDL-CAD | 0.3402 | 1.17E-45 | 0.31 (0.24, 0.38) | 0.2014 | 8.56E-27 |
| BMI-Edu | -0.2241 | 3.74E-14 | -0.12 (-0.14, -0.11) | -0.1892 | 6.15E-35 |
| SHeight-BMI | -0.1278 | 1.40E-22 | -0.13 (-0.14, -0.11) | -0.0854 | 9.01E-23 |
| SBP-BWeight | -0.2565 | 9.85E-08 | -0.13 (-0.16, -0.1) | -0.1646 | 1.20E-11 |
| SBP-SHeight | -0.3657 | 4.81E-08 | -0.12 (-0.15, -0.1) | -0.0967 | 0.004422636 |
| SHeight-SBP | -0.0759 | 5.74E-05 | -0.08 (-0.09, -0.07) | -0.0652 | 1.25E-15 |
| SHeight-SVstat | -0.0465 | 4.76E-09 | -0.01 (-0.02, -0.01) | -0.0328 | 6.78E-12 |
| BMI-HDL | -0.3760 | 3.54E-56 | -0.28 (-0.29, -0.26) | -0.3630 | 3.17E-111 |
| SHeight-LDL | -0.0716 | 4.26E-09 | -0.04 (-0.05, -0.02) | -0.0298 | 5.07E-06 |
| BWeight-CAD | -0.1745 | 2.05E-06 | -0.21 (-0.28, -0.14) | -0.0978 | 2.83E-05 |
| SHeight-CAD | -0.0802 | 3.72E-20 | -0.15 (-0.18, -0.12) | -0.0482 | 2.18E-12 |
| HDL-CAD | -0.1729 | 7.00E-31 | -0.26 (-0.3, -0.21) | -0.0778 | 5.45E-10 |

**Table S8: Table comparing the causal estimates of LHC-MR, CAUSE, and IVW for trait pairs that had a significant causal effect in LHC-MR and CAUSE**. The column showing the gamma (causal effect) estimate of the CAUSE method also reports its 95% credible intervals. A complete table for all the studied pairs is found in the Supplementary Table S5.
